# From dysphoria to anhedonia: Age-related shift in the link between cognitive and affective symptoms

**DOI:** 10.1101/2025.03.26.25324666

**Authors:** Daniel Harlev, Aya Vituri, Moni Shahar, Noham Wolpe

**Affiliations:** Department of Physical Therapy, The Stanley Steer School of Health Professions, Gray Faculty of Medical & Health Sciences, Tel Aviv University, George Wise St. 30, Tel Aviv 6997801, Israel; Department of Psychiatry, Rambam Health Care Campus, HaAliya HaShniya St 8, Haifa 3109601, Israel; Tel Aviv Center for Artificial Intelligence & Data Science (TAD), Tel Aviv University, George Wise St. 30, Tel Aviv 6997801, Israel; Sagol School of Neuroscience, Tel Aviv University, George Wise St. 30, Tel Aviv 6997801, Israel

**Keywords:** Aging, Depression, Bridging, Cognition, Gray matter volume, Symptoms

## Abstract

**Background:** Depression in aging shows heterogeneous symptoms across cognitive, affective, and neurobiological domains. Traditional categorical diagnoses may not capture these complex patterns, prompting a shift toward dimensional or domain-based approaches. We examined whether the symptoms that bridge cognition and affect differ by age, and explored their associations with brain structure.

**Methods:** Data from 756 young (≤45 years) and 1230 older (≥65 years) adults from the Cambridge Centre for Ageing and Neuroscience were analysed. Cognition was assessed using the Addenbrooke’s Cognitive Examination Revised, and depressive and anxiety symptoms with the Hospital Anxiety and Depression Scale. Graphical LASSO was used to construct cognitive-affective networks, testing for age-related differences in strength and bridging centrality measures. Building on these findings, we further examined the association between bridging symptoms, cognition, and gray matter volume (GMV).

**Results:** Symptom strength centrality was similar across groups. However, in older but not in younger adults, depressive symptoms were more strongly connected to cognitive symptoms than anxiety symptoms. The primary bridging symptom shifted with age, from dysphoria in young adults to anhedonia in older adults. Follow-up analyses indicated that the anhedonia-to-dysphoria difference was associated with the relationship between GMV and cognition, particularly in older adults.

**Conclusions:** Cognitive-affective bridging symptoms differ with age, with anhedonia replacing dysphoria as the key bridge in older adults. This shift was linked to differences in how GMV relates to cognition in late life. These results highlight the need to target different symptoms to alleviate cognitive-affective manifestations across the lifespan.

## Introduction

Traditional psychiatric diagnosis relies on categorical classifications, assigning diverse symptoms to single diagnostic entities such as Major Depressive Disorder (MDD)^1^. However, evidence increasingly shows that depressive disorders are highly heterogeneous: patients with the same diagnosis may present with very different symptom profiles, severities, and functional impairments ^2,3^. Such heterogeneity challenges the validity of categorical diagnoses and raises questions about their utility for understanding mechanisms, predicting outcomes, or guiding personalized treatment ^4^.

These diagnostic challenges become even more pronounced in the context of aging. Older adults experience significant changes across cognitive, affective, and neurobiological domains, and mental health diagnoses such as depression and anxiety are common in this population ^5–7^. Importantly, depressive and anxiety symptoms in older adults often overlap with cognitive decline, both clinically and biologically, making it difficult to disentangle psychiatric syndromes from neurodegenerative processes ^8–10^. This overlap highlights the limitations of categorical thinking, as symptoms may span multiple domains such as apathy or anhedonia, and carry different implications depending on the individual ^11–13^. Approaches that move beyond categorical labels toward understanding how specific symptoms interrelate—such as network analysis—may offer clearer insights into the complex mental health landscape in older age^14^.

Symptom network analyses offer a powerful approach for understanding how specific symptoms interact and cluster into meaningful patterns across domain^15,16^. However, most existing studies have focused either on younger adults or on older adults in isolation, without comparing networks across the lifespan^14,17,18^. Moreover, many have examined either cognitive or affective domains separately, rather than investigating how these domains interact. In older adults, previous studies have focused on affective symptoms, while excluding cognitive domains ^19,20^, and the few studies combining affective and cognitive domains had methodological limitations. For example, one study ^21^ used less sensitive cognitive measures like the Mini-Mental State Examination, which lacks sensitivity to specific cognitive domains such as executive function, and is particularly limited in detecting mild cognitive impairment (MCI), a condition highly relevant in aging populations and often overlapping with affective symptoms^22^.

Another study^23^ focused on clinical populations, limiting broader applicability to the general population. Finally, few studies have integrated neurobiological measures, such as gray matter volume (GMV), which are essential for linking symptom networks to underlying brain structure and understanding potential mechanisms driving cognitive-affective interactions.

Here, we address these gaps by directly comparing cognitive-affective networks in the general population across the lifespan. We analyzed data from the Cambridge Centre for Ageing and Neuroscience, which is a population-based cohort of overall healthy individuals across the lifespan. Cognitive performance was assessed with the Addenbrooke’s Cognitive Examination Revised (ACE-R) to assess performance in key cognitive domains. Affective symptoms were assessed with the Hospital Anxiety and Depression Scale (HADS). We compared strength centrality to assess overall network structure and bridge centrality measures to assess the cognitive-affective links in both young and older adults. We further tested whether depression and anxiety symptoms differentially contribute to linking cognitive and affective symptoms across age groups. Lastly, we explored whether the age-related difference in bridging symptoms moderates the relationship between GMV and cognitive performance across the lifespan.

## Methods

### Participants

We analyzed data from the Cambridge Centre for Ageing and Neuroscience (Cam-CAN)—a large-scale project focusing on aging ^24^. Cam-CAN recruited a population-based sample of approximately 2600 adults aged 18 years and over from the general population via Primary Care Trust’s lists within the Cambridge City area in the UK. Ethical approval for the study was granted by the Cambridgeshire 2 Research Ethics Committee (reference: 10/H0308/50), and written informed consent was obtained from all participants prior to their participation. The dataset includes detailed demographic, cognitive, and neuroimaging data.

Participants were categorized into two age groups: young (≤45 years) and older (≥65 years), to test for age-related differences in cognitive-affective networks. Cognitive performance was assessed using the Addenbrooke’s Cognitive Examination-Revised (ACE-R) ^25^, which evaluates five domains: memory, fluency, language, visuospatial abilities, and orientation. Affective symptoms were measured using the Hospital Anxiety and Depression Scale (HADS) ^26^, which includes subscales for depressive (HD1–HD7) and anxiety (HA1–HA7) symptoms. After excluding participants with missing demographic or clinical data, the groups included 756 young adults and 1230 older adults.

### Network Analyses

Sparsity-based networks were constructed separately for each age group using the Graphical LASSO method, as done previously ^27^. We included participants across the full range of HADS scores, including those with low or zero scores, to capture the entire spectrum of affective symptomatology and its relationship with cognitive performance, as done previously ^21^. The use of Graphical LASSO allowed us to include all participants by penalizing weak or noisy connections to ensure network construction was robust across the full range of affective and cognitive scores. Non-significant connections were removed based on the Extended Bayesian Information Criterion (EBIC) ^28^ for model selection. Nodes represented cognitive and affective domains, with edges reflecting regression coefficients. Covariates, including sex and education (number of years in education), were included to account for potentially confounding effects. All non-categorical data were Z-scored before entry into the analysis.

In line with our study goals and with previous research, we computed two key metrics in the networks ^29,30^. First, we sought to identify the importance of each node within the whole network by calculating strength centrality. Strength centrality quantifies the total connectivity of a node by summing the absolute weights of all edges connected to it. Higher strength centrality indicates that a node is more integrated within the network, i.e., it has stronger connections to multiple other nodes. This allowed us to test whether overall network structure remained similar with age, with similarly important nodes.

Second, we sought to identify the nodes linking cognitive and affective symptoms by computing two independent and complementary centrality measures. Betweenness centrality quantifies how often a node appears on the shortest paths between pairs of other nodes in a network (where shortest path is the minimum number of steps/lowest-weight sum needed to travel between two nodes in a network). In other words, betweenness centrality reflects a node’s role as an intermediary. When a node mediates a large proportion of the shortest paths, it plays a critical role in connecting otherwise separate regions of the network, and is thus interpreted as a bridging measure ^31^. To complement this approach, we also computed Bridge Expected Influence (BEI), which measures a node’s connections spanning different pre-defined communities, weighted by their influence on neighboring nodes ^30^. Unlike betweenness centrality, BEI is domain-sensitive and requires *a priori* labelling of the different symptom domains (cognitive and affective).

We first assessed the consistency of centrality ranks across age groups using Spearman (rank) correlation to determine whether the overall structure of centrality values remained similar between young and older adults. Next, we reported the nodes with the highest centrality value in each domain (depression, anxiety and cognition) for each age groups to identify possible age-related differences in the importance of network nodes. Finally, we tested for differences in centrality measures between depression and anxiety nodes in each group. This tests whether within each group, there was any significant differences in centrality measures between depression and anxiety symptoms. To test this, permutation testing with 5,000 iterations was used within each age group to compare centrality measures between depression-related (HD1–HD7) and anxiety-related (HA1–HA7) nodes. The test was performed by randomly shuffling the depression and anxiety labels while maintaining the original network topology, generating a ‘null’ distribution of expected differences by chance alone. Statistical significance was determined as the proportion of permutations in which the shuffled difference exceeded the observed value (with *p* < 0.05 considered statistically significant).

### *Post hoc* GLM Analyses

Building on the network analyses, we conducted exploratory post hoc Generalized Linear Models (GLMs) to test whether the age-related difference in bridging symptoms modulated the relationship between GMV and cognition. While previous research has shown that depressive symptoms moderate GMV-cognition associations ^32^, these studies did not examine how specific depressive symptoms contribute to this relationship.

The analysis included 733 participants in total, who took part in Cam-CAN Stage 2 and Cam-CAN Frail, which followed similar neuroimaging protocols in individuals with lower scores in the ACE-R ^24,33^. Structural MRI data were acquired using a 3T Siemens TIM Trio scanner with T1-weighted MPRAGE sequences (TR = 2250 ms, TE = 2.99 ms, TI = 900 ms, flip angle = 9°, isotropic voxel size = 1 mm³) ^34^. Preprocessing followed standard protocols in SPM12, including segmentation into gray matter. To account for individual differences in head size, total intracranial volume (TIV) was computed from the segmentation, and GMV was included in the model as the ratio GMV/TIV.

We examined whether anhedonia versus dysphoria moderated the relationship between GMV and cognitive performance. To quantify this, we computed the anhedonia-to-dysphoria difference(HD7−HD3), where higher values indicate a greater relative severity of anhedonia compared to dysphoria ^35^.

The GLM included cognitive performance (total ACE-R score) as the dependent variable. Age, GMV and anhedonia-to-dysphoria difference were the independent variables. Two-way interactions were tested to determine whether anhedonia-to-dysphoria difference moderated the relationship between GMV and cognition across age groups. For consistency with the network analysis, education and sex were included as covariates, and LASSO regularization was implemented. To complement the model results. All statistical analyses were conducted in Python (v3.11.5) ^36^.

## Results

### Participants and item severity

Participant demographics are summarized in Table 1 for both young and older adult groups. Sex distribution was similar across groups, but education level was significantly higher for younger compared to older adults. Depressive symptoms varied between the two groups. Older adults had significantly higher scores on items related to psychomotor slowing and reduced anticipatory anhedonia/enjoyment, while young adults scored higher on measures of dysphoria/cheerfulness and consummatory anhedonia/enjoyment. There were no significant differences between groups for apathy/interest in appearance or laughter. In contrast, anxiety symptoms were consistently higher among young adults across all items. Lastly, young adults had better cognitive performance compared to older adults across all cognitive domains.

**Table 1.** Summary of demographics and item-wise comparisons between young and older adults. Demographic, cognitive, and affective items for young and older adults are presented. Education was categorized according to the English system: “None” (no education beyond age 16), “GCSE” (General Certificate of Secondary Education), “A Levels” (General Certificate of Education Advanced Level), and “University” (undergraduate or graduate degree). Continuous variables, including age, and cognitive and affective measures, are reported as mean ± standard deviation and compared using independent t-tests. Categorical variables, such as sex distribution and education, are expressed as percentages and compared with chi-square tests. All *p-values* were corrected for multiple comparisons using FDR correction.

| Variable | Young Adults<br>(mean ± SD<br>or %) | Older Adults<br>(mean ± SD<br>or %) | Test Statistic | p-value (FDR<br>corrected) |
| --- | --- | --- | --- | --- |
| **Age** | 33.49 ± 6.87 | 79.29 ± 7.41 | | |
| **Sex** | 42.9% | 43.5% | 1.26 | 0.557 |
| **Education** | None: 1.7%;<br>GCSE: 4.1%;<br>A Level: 8.1%;<br>University:<br>86.1% | None: 29.5%;<br>GCSE: 7.7%;<br>A Level 3.1%;<br>University:<br>59.7% | 348.68 | <0.001 |
| **Anhedonia/Enjoyment**<br>**(HD1)** | 0.46 ± 0.67 | 0.61 ± 0.78 | -4.65 | <0.001 |
| **Laughter (HD2)** | 0.26 ± 0.50 | 0.27 ± 0.54 | -0.25 | 0.802 |
| **Dysphoria/Cheerful**<br>**Feeling (HD3)** | 0.35 ± 0.54 | 0.27 ± 0.52 | 3.11 | 0.002 |
| **Psychomotor**<br>**retardation/Slowed**<br>**Down (HD4)** | 0.70 ± 0.72 | 1.36 ± 0.93 | -16.47 | <0.001 |
| **Apathy/Loss of**<br>**interest in Appearance**<br>**(HD5)** | 0.50 ± 0.74 | 0.47 ± 0.71 | 0.85 | 0.434 |
| **Anticipatory**<br>**anhedonia/Look**<br>**Forward to Things**<br>**(HD6)** | 0.37 ± 0.65 | 0.53 ± 0.72 | -4.91 | <0.001 |
| **Anhedonia/Enjoyment**<br>**of Book/Media (HD7)** | 0.28 ± 0.63 | 0.19 ± 0.52 | 3.52 | <0.001 |
| **Feeling of Tension**<br>**(HA1)** | 1.06 ± 0.65 | 0.78 ± 0.62 | 9.52 | <0.001 |
| **Frightened Feeling** | 0.69 ± 0.81 | 0.53 ± 0.74 | 4.38 | <0.001 |
| **(HA2)** |  |  |  |  |
| **Worrying Thoughts (HA3)** | 0.98 ± 0.87 | 0.76 ± 0.80 | 5.86 | <0.001 |
| **Relaxed Feeling (HA4)** | 0.84 ± 0.68 | 0.60 ± 0.63 | 7.93 | <0.001 |
| **Butterflies in Stomach (HA5)** | 0.61 ± 0.64 | 0.43 ± 0.59 | 6.49 | <0.001 |
| **Restless Feeling (HA6)** | 1.21 ± 0.90 | 0.92 ± 0.82 | 7.36 | <0.001 |
| **Feeling of Panic (HA7)** | 0.54 ± 0.67 | 0.49 ± 0.65 | 1.68 | 0.107 |
| **Memory** | 0.93 ± 0.10 | 0.82 ± 0.17 | 15.81 | <0.001 |
| **Fluency** | 0.89 ± 0.13 | 0.78 ± 0.19 | 15.04 | <0.001 |
| **Language** | 0.94 ± 0.10 | 0.92 ± 0.10 | 3.65 | <0.001 |
| **Visuospatial** | 0.98 ± 0.05 | 0.91 ± 0.14 | 13.52 | <0.001 |
| **Orientation** | 0.97 ± 0.06 | 0.93 ± 0.10 | 10.15 | <0.001 |

### Network Analyses

Cognitive-affective networks for young and older adults were constructed for young and older adults. In both age groups, nodes clustered into cognitive and affective domains, with predominantly positive connections within each domain, and largely negative connections between them, although some inter-domain connections were positive (Fig. 1).

**Figure 1.** Sparsity-based networks for young and older adults. A) Cognitive-affective symptom network for young adults. Each Node represents a cognitive (blue) or affective (red) item, and edges representing conditional relationships estimated via the Graphical LASSO method. Green edges indicate positive relationships, and orange edges indicate negative relationships. Edge thickness corresponds to the strength of the relationships. B) Same as (A) but for older adults.

To examine the network structure and whether it differs between age groups, we calculated two key centrality measures, namely strength centrality and betweenness centrality. First, strength centrality, which quantifies the overall importance of a node by summing up the overall connection weight of a node, was computed. The pattern of strength centrality values was overall consistent between groups (Fig. 2A). The rank of strength centrality values was indeed highly correlated between young and older adults (Spearman’s ρ = 0.795, *p =* 4.80e-05). In terms of their values, the strongest cognitive nodes for both young and older adults were memory (young: 0.848, old: 0.731), fluency (young: 0.694, old: 0.883) and language (young: 1.063, old: 0.991). Among the affective nodes, the nodes with highest strength centrality for both groups were anticipatory anhedonia/enjoyment (HD6) (young: 1.045, old: 1.155) for depression, and worrying thoughts (HA3) (young: 1.031, old: 1.070) for anxiety. A permutation test (n = 5000) comparing mean strength centrality between depression-related nodes (HD1–HD7) and anxiety-related nodes (HA1–HA7) showed no significant differences in either age group (young: mean observed difference = −0.217, *p =* 0.618; old: mean observed difference = −0.207, *p =* 0.531). Together, these results suggest that overall network structure was similar between young and older adults.

**Figure 2.** Centrality measures across age groups. A) Strength centrality for young (purple) and older adults (green). Strength centrality represents the overall connectivity of each node, calculated as the sum of absolute edge weights connected to the node. Higher values indicate nodes with greater influence across the network. The centrality measures were derived from sparsity-based networks, calculated using the Graphical LASSO method with extended Bayesian Information Criteria thresholding. The x-axis represents centrality values, and the y-axis lists the nodes. B) Same as (A), but for betweenness centrality for young (purple) and older adults (green). Betweenness centrality represents the importance of a node in a network based on how often it appears on the shortest paths between other nodes. A node with high betweenness centrality acts as a key connector within the network.

Next, we computed two independent and complementary bridging measures. First, we computed each node’s betweenness centrality, which quantifies how often a node appears on the shortest paths between other nodes. In contrast to strength centrality, betweenness centrality pattern was different between young and older adults (Fig. 2B). The rank of betweenness centrality values was not correlated between young and older adults (Spearman’s ρ = 0.330, *p* = 0.168). In terms of their value, the symptoms with the highest betweenness value were dysphoria (HD3) in young adults (0.4575) but anhedonia (HD7) in older adults (0.2843). A permutation test (n = 5000) comparing mean betweenness centrality between depression- and anxiety-related nodes showed group-specific differences. Specifically, in young adults, depression and anxiety nodes did not differ significantly (observed difference = 0.053, *p =* 0.702). However, in older adults, betweenness centrality was significantly higher for depression-related nodes compared to anxiety-related nodes (observed difference = 0.326, *p =* 0.033).

The second bridging measure we computed was BEI, which quantifies bridging based on predefined node (symptom) labels. The analysis showed similar results to those of betweenness centrality above (Fig. S1), namely no rank correlation between BEI in young and older adults (Spearman’s ρ = 0.326, *p* = 0.173); dysphoria (BEI = −0.0741) and anhedonia (BEI = −0.2012) had the highest negative BEI values in young and older adults, respectively; and no differences in young adults (mean observed difference = −0.071, *p =* 0.548) but significantly higher depression-related BEI compared to anxiety-related BEI in older adults (mean observed difference = −0.443, *p =* 0.0014). Interestingly, rumination (HA3) showed a surprisingly positive BEI value in both young (0.0314) and older adults (0.0516)

### Bridging symptoms, GMV, and cognitive performance

To examine whether the age-related difference in bridging symptoms moderates the relationship between GMV and cognitive performance, we conducted a *post hoc* GLM analysis (Table 2). For the main contrasts of interest, significant effects were observed for the anhedonia-to-dysphoria difference × GMV and anhedonia-to-dysphoria difference × age, indicating that more anhedonia relative to dysphoria was associated with a more positive relationship between GMV and cognition. However, the three-way interaction (anhedonia-to-dysphoria difference × GMV × age) was not significant.

**Table 2.** Generalized Linear Model (GLM) results for predictors of ACE-R. Results from a Generalized Linear Model (GLM) predicting cognitive performance (ACE-R) based on anhedonia-to-dysphoria difference (calculated as (HD7-HD3), age, GMV (calculated as GMV-to-TIV ratio), and their interactions. All continuous variables were z-scored prior to model entry. The model includes main effects for anhedonia-to-dysphoria difference, age, GMV, sex, and education, as well as two-way and three-way interaction terms. Significance levels: *p < 0.05, **p < 0.01, ***p < 0.001. Model fit: R² = 0.319, Adjusted R² = 0.310.

| **Predictor** | **β estimate** | **SE** | **t-statistic** | **p-value** | **95% CI**<br>**(Lower)** | **95% CI**<br>**(Upper)** |
| --- | --- | --- | --- | --- | --- | --- |
| **Intercept** | -0.6187 | 0.101 | -6.149 | 1.29e-09 | -0.816 | -0.421 |
| **Anhedonia**<br>**to**<br>**dysphoria**<br>**difference)** | -0.0453 | 0.041 | -1.106 | 0.269 | -0.126 | 0.035 |
| **Age** | -0.2496 | 0.055 | -4.530 | 6.90e-06<br>*** | -0.358 | -0.141 |
| **GMV** | 0.1274 | 0.056 | 2.283 | 0.023 * | 0.018 | 0.237 |
| **Anhedonia-**<br>**to-**<br>**dysphoria**<br>**difference×**<br>**GMV** | 0.1329 | 0.056 | 2.377 | 0.018 * | 0.023 | 0.243 |
| **Anhedonia-**<br>**to-**<br>**dysphoria**<br>**difference ×**<br>**Age** | 0.1198 | 0.057 | 2.108 | 0.035 * | 0.008 | 0.231 |
| **GMV × Age** | 0.1858 | 0.033 | 5.678 | 1.98e-08<br>*** | 0.122 | 0.250 |
| **anhedonia-**<br>**to-dysphoria**<br>**difference×**<br>**GMV × Age** | -0.0069 | 0.028 | -0.248 | 0.804 | -0.061 | 0.048 |
| **Sex** | -0.0300 | 0.065 | -0.461 | 0.645 | -0.158 | 0.098 |
| **Education** | 0.3062 | 0.033 | 9.171 | 4.80e-19<br>*** | 0.241 | 0.372 |

Although the three-way interaction was not statistically significant, suggesting that the relationship between GMV and cognition did not differ systematically across all levels of age (age included in the GLM as continuous variable), exploratory subgroup analyses revealed additional meaningful differences. Specifically, in young adults (≤ 45 years old), the correlation between GMV and cognitive performance was not significant for both people with higher anhedonia relative to dysphoria (*r* = 0.030, *p* = 0.738) and people with higher dysphoria relative to anhedonia (*r* = −0.256, *p* = 0.151) (Fig. 3A). In contrast, in older adults (≥ 65 years old) with higher anhedonia relative to dysphoria, GMV was significantly positively correlated with cognitive performance. (*r* = 0.333, *p =* 8.606e-05), but not for older adults with higher dysphoria relative to anhedonia (*r* = 0.150, *p* = 0.515) (Fig. 3B). The correlation between GMV and cognition was indeed significantly stronger in older compared to young adults for individuals with high anhedonia relative to dysphoria (*z* = −2.502, *p* = 0.012), but not for individuals with higher dysphoria relative to anhedonia (*z* = −1.386, *p* = 0.166). These findings indicate that while the full three-way interaction may lack consistency to reach statistical significance, subgroup-specific associations—particularly in older adults with more prominent anhedonia—may still be present.

**Figure 3.** Associations between cognitive performance and gray matter volume as a function of anhedonia/dysphoria by age. A) Scatter plot showing the relationship between cognitive performance, measured with the Addenbrooke’s Cognitive Examination Score – Revised (ACE-R) and gray matter volume (GMV) for younger adults (≤ 45 years old). Gray matter volume is shown as a proportion of total intracranial volume. For illustration, data points are plotted separately for individuals with high anhedonia relative dysphoria (blue) and high dysphoria relative to anhedonia (red), based on a median split. In the statistical model, both age and anhedonia-to-dysphoria difference (HD7 - HD3) were analyzed as continuous variables. Regression lines represent fitted trends for each group, with shaded areas indicating 95% confidence intervals. B) Same as (A) but for older adults (≥ 65 years).

## Discussion

Our study investigated the structure of cognitive-affective symptom network across the lifespan. Overall network structure was similar across groups, Importantly, however, distinct patterns of symptom connectivity emerged for the bridging of affective and cognitive symptoms between age groups. Across two independent bridging measures, dysphoria was the strongest cognitive-affective bridging symptom in young adults, while anhedonia was the strongest bridging symptom for older adults. Moreover, depressive symptoms had significantly higher cognitive-affective bridging values compared to anxiety symptoms, specifically for older adults. Moreover, higher anhedonia-to-dysphoria difference was associated with stronger and more positive relationship between GMV and cognition, with this effect being most pronounced in older adults. These findings suggest an age-related shift in the symptoms that bridge cognitive and affective symptoms.

### Model free symptom severity

Cognitive and anxiety items showed a distinct age-related pattern, with anxiety symptoms being more severe in younger adults, while cognitive function was better preserved in this group compared to older adults.

This pattern reflects the generally higher prevalence of anxiety in younger populations within the general population, consistent with prior epidemiological findings ^37,38^. In contrast, depressive symptoms exhibited greater variability, consistent with the large heterogeneity of depression manifestation ^2,3^. Older adults reported more severe psychomotor slowing, while younger adults reported more severe mood-related symptoms. These findings are consistent with known age-related differences in depressive symptom profiles ^39,40^, and our principal aim was to examine cognitive-affective interactions by age using symptom network analyses.

### Maintenance of overall cognitive-affective network structure

Strength centrality, which reflects the overall connectivity of nodes, showed remarkable stability across age groups. Within the cognitive nodes, memory and language consistently retained high centrality values in both young and older adults. This likely reflects their role as core cognitive functions that remain relevant regardless of age. Similarly, within the affective nodes, anticipatory anhedonia (HD6) and worrying thoughts (HA3) retained high centrality values across age groups. This pattern aligns with previous findings indicating that the core organization of depressive and anxiety symptom networks, including the centrality of key symptoms, remains largely stable across the lifespan ^14^.

This cross-domain consistency across the lifespan supports the view that affect and cognition form an integrated clinical construct that is preserved with age. Indeed, meta-analytic evidence confirms that depressive symptoms are reliably associated with cognitive control deficits from childhood through old age ^41^. This stability suggests that depression, anxiety, and cognitive symptoms belong to a common clinical syndrome that remains consistent across age. But while the structure appears similar, the mechanisms linking these domains may change with age—pointing to potentially distinct pathways.

### Age-related shift in cognitive-affective bridging symptoms

Using two independent and complementary bridging centrality measures (betweenness and BEI), we identified a difference in how affective symptoms integrate with cognition across age groups. In younger adults, mood-related symptoms, and particularly cheerful feelings, were strongly connected to cognitive nodes, reflecting the importance of emotional positivity in cognitive performance during earlier life stages ^42^. In contrast, in older adults, anhedonia became the principal bridging symptom, reflecting its stronger association with cognitive deficits in late life ^1^.

A shift from mood-related link to cognitive performance in younger adults to anhedonia-related link to cognitive performance in older adults may reflect age-related neurobiological changes in affective regulation. Whereas younger adults show strong reciprocal links between mood and cognitive function ^43^, aging is associated with reduced emotional responsivity but increased motivational deficits ^11^. Consistent with this, in Late-Life Depression (LLD), apathy—but not dysphoria—exhibits the strongest association with cognitive dysfunction ^12^.

Building on these results, we conducted a GLM linking cognition with anhedonia-to-dysphoria, GMV and age. While the three-way interaction between anhedonia-to-dysphoria, GMV, and age was not significant, post hoc analyses revealed specific patterns that merit attention. Specifically, a positive association between GMV and cognition was observed in older adults with greater anhedonia relative to dysphoria. This effect was not evident in young adults or in individuals with relatively greater dysphoria. The absence of a significant three-way interaction suggests that these effects do not generalize across all subgroups or may not follow a consistent interaction pattern across the full range of the sample. Nonetheless, the subgroup-specific findings suggest that the relationship between brain structure and cognitive performance may become more tightly coupled in certain clinical or demographic contexts—such as older individuals experiencing prominent anhedonia.

These results are consistent with observations that in older adults, anhedonia is more closely linked to cognitive decline and structural brain changes ^44^, while in young adults, emotional reactivity changes are more closely linked to cognitive ^45^. One possible explanation is cognitive reserve, which reflects individual differences in the brain’s ability to maintain function despite structural decline ^46^. Notably, the stronger association between cognition and GMV was seen particularly in older adults. The stronger association between lower GMV and poorer cognitive performance in older individuals with high anhedonia can thus reflect reduced cognitive reserve. As anhedonia increases, cognition may become more dependent on brain integrity ^47,48^.

Beyond structural differences, network-based measures further highlight this shift in cognitive-affective interactions with aging. While overall network strength remained comparable between depressive and anxiety symptoms, depressive symptoms in older adults exhibited stronger connectivity to cognitive nodes compared to anxiety symptoms. This finding underscores the increasingly tight link between depressive symptoms and cognitive functioning in older populations ^49,50^.

Together, these findings suggest that in aging, depression—particularly its anhedonic aspects—may be increasingly intertwined with cognitive decline and underlying brain vulnerability. Indeed, apathy and anhedonia have been increasingly recognized as key early features of neurodegenerative conditions, including Alzheimer’s disease, Parkinson’s disease, and frontotemporal dementia (Steffens et al., 2022). Their presence in these disorders reflects disruptions in reward-processing circuits, which are crucial for both cognitive and affective regulation ^11^. In aging, anhedonia may represent a distinct mechanism of depression—one more closely tied to neurodegenerative pathways than to mood dysregulation.

### Clinical implications: rethinking late-life depression treatments

If anhedonia reflects a different mechanism of depression in aging, which is linked to neurodegeneration rather than traditional mood disturbances, then current treatment approaches may need to be reconsidered. Currently, depression in older adults is treated similarly to depression in younger adults, despite lower efficacy ^8^. While serotonin-based antidepressants remain standard, anhedonia and apathy may be more strongly linked to dysfunction in dopamine-mediated reward circuits ^10^. Dopaminergic interventions, cognitive-behavioral strategies emphasizing goal-directed activity, and neurostimulation approaches have shown promise in targeting motivational impairments rather than mood dysregulation alone ^51^. Future research should investigate whether such approaches can prove efficacious in older adults suffering from anhedonic depression.

### Strengths and limitations

To our knowledge, our study is the first to directly compare differences in cognitive and affective symptoms in a large population-based study. The results from multiple centrality measures converged to show both stable and dynamic patterns in cognitive-affective network. Moreover, by integrating neuroimaging data, the analysis provided supplementary evidence for a possible biological link underlying cognitive-affective interactions with age.

However, several limitations must be considered. First, the cross-sectional design precludes causal inferences about the observed shifts in network patterns. Second, our neuroimaging analyses focused on global GMV rather than region-specific effects, providing a standardized clinical measure, but limiting insights into the role of specific brain structures in cognitive-affective interactions. Third, the study did not include a clinical population, which restricts the generalizability of findings to clinical settings. However, we argue this may better reflect patterns in the general population. Fourth, reliance on self-reported symptom measures could introduce biases, particularly in older adults, which would affect the accuracy of symptom network measures. This limitation is further highlighted by the interpretation of single-item measures, which may not fully capture the complexity of syndromes like anhedonia and apathy. This oversimplification, while common to many other symptom network studies, calls for caution when interpreting the results of these studies.

## Conclusion

Depressive symptoms, and particularly anhedonia, become more central in linking cognitive and affective symptoms with age. Higher anhedonia relative dysphoria was associated with stronger and more positive relationship between GMV and cognition, particularly in older adults. These findings highlight how aging alters cognitive-affective interactions and underscore the need for therapeutic strategies that account for both symptom profiles and neurobiological factors across the lifespan.

## Data and code availability

All data used for this work are publicly available upon signing data sharing agreement on https://cam-can.mrc-cbu.cam.ac.uk/dataset/. Code used to analyse the data and generate the figures is available on https://github.com/dharlev/From-dysphoria-to-anhedonia-Age-related-shift-in-the-link-between-cognitive-and-affective-symptoms.

## Consent Statement

All participants provided written informed consent prior to participation. Ethical approval was obtained from the Cambridgeshire 2 Research Ethics Committee (reference: 10/H0308/50).

## Author Contributions

DH designed the study, conducted the analysis, and drafted the manuscript. AV assisted with data preprocessing and contributed to result interpretation. MS contributed to the conceptual framing and data preprocessing. NW supervised the project and provided critical feedback throughout all stages.

## Acknowledgements

Cam-CAN research was supported by the Biotechnology and Biological Sciences Research Council (BB/H008217/1). DH was supported by an Israel Science Foundation Mavri fellowship. NW was supported by an Israel Science Foundation Personal Research Grant (1603/22). We are grateful to the Cam-CAN respondents and their primary care teams in Cambridge for their participation in this study.

